# (Re-)modelling of the disease and mortality burden of the 1918-1920 influenza pandemic in Zurich, Switzerland

**DOI:** 10.1101/2024.03.14.24304276

**Authors:** Ella Ziegler, Katarina L. Matthes, Peter W. Middelkamp, Verena Schünemann, Christian L. Althaus, Frank Rühli, Kaspar Staub

## Abstract

**Background:** Our study aims to enhance future pandemic preparedness by integrating lessons from historical pandemics, focusing on the multidimensional analysis of past outbreaks. It addresses the gap in existing modelling studies by combining various pandemic parameters in a comprehensive setting. Using Zurich as a case study, we seek a deeper understanding of pandemic dynamics to inform future scenarios.

**Data and methods:** We use newly digitized weekly aggregated epidemic/pandemic time series (incidence, hospitalisations, mortality and sickness absences from work) to re-model the 1918-1920 pandemic in Zurich and investigate how different parameters correspond, how transmissibility changed during the different waves, and how public health interventions were associated with changes in these pandemic parameters.

**Results:** In general, the various time series show a good temporal correspondence, but differences in their expression can also be observed. The first wave in the summer of 1918 did lead to illness, absence from work and hospitalisations, but to a lesser extent to increased mortality. In contrast, the second, longest and strongest wave in the autumn/winter of 1918 also led to greatly increased (excess) mortality in addition to the burden of illness. The later wave in the first months of 1920 was again associated with an increase in all pandemic parameters. Furthermore, we can see that public health measures such as bans on gatherings and school closures were associated with a decrease in the course of the pandemic, while the lifting or non-compliance with these measures was associated with an increase of reported cases.

**Discussion:** Our study emphasizes the need to analyse a pandemic’s disease burden comprehensively, beyond mortality. It highlights the importance of considering incidence, hospitalizations, and work absences as distinct but related aspects of disease impact. This approach reveals the nuanced dynamics of a pandemic, especially crucial during multi-wave outbreaks.

## 1. Introduction

Integrating the experience and lessons learned from historical pandemics is not only beneficial during an ongoing pandemic outbreak ^1–7^ but can also help inform possible scenarios for future pandemic planning and preparedness ^5,8–13^. However, signature features of past pandemics need to be researched ^14^ and these experiences need to be communicated, and ultimately adapted to new challenges and contexts. And this is where Switzerland, like other European countries, has catching up to do, as the absence of strong mortality effects of the pandemics from 1957 onwards resulted in the experience gained and the immediate risk awareness being forgotten over many years ^15^.

The most extensively researched past influenza outbreak is the 1918-1920 influenza pandemic (“Spanish flu”), which caused 20-100 Mio. deaths worldwide and was therefore considered as unusually deadly ^16,17^. Most existing studies focus on mortality data and thus on the worst possible outcome of a pandemic. But it is the nature of pandemics triggered by respiratory infections that most of the population becomes infected or ill, and the vast majority of those infected survive the disease. Thus, if one wants to describe the health burden of a pandemic more comprehensively than with deaths, one must also include morbidity parameters (e.g., incidence, hospitalizations, absences from work due to illness). There are few studies in the literature that link several of these pandemic parameters to provide a more holistic picture of the course of the 1918-1920 pandemic. With our contribution we aim to demonstrate the added value of such a multi-perspective approach. In addition, COVID-19 reminded us of the specific challenges related to multi-wave pandemics. In this respect, there is hardly any comparison other than the 1918-1920 pandemic.

By European comparison, Switzerland was hit medium-hard by the 1918-1920 pandemic ^18^. In Switzerland, the 1918-1920 pandemic occurred in three to four waves, causing at least 2 million infections (at least two-thirds of the population) and about 25,000 deaths (0.67% of the population) between summer 1918 and spring 1920 ^19^. As elsewhere, the death toll included a disproportionate number of young people and especially men. No less than ca. 60% of all deaths in Switzerland come from the age segment that is usually most resistant to infectious diseases. Therefore, the 1918 influenza pandemic is considered the greatest demographic catastrophe of the 20th century in Switzerland ^19,20^. Recent research on Switzerland can be divided into a) national descriptions based on mortality figures ^19,20^, b) qualitative reconstructions of the epidemic in selected member states (cantons) ^19,21–24^, and c) a growing number of quantitative studies modelling the pandemic based on hospitalisation data in Geneva ^25,26^ and incidence data from Bern ^27,28^.

The modelling studies, however, lack a case study for which various pandemic parameters are brought together in a multi-dimensional setting to gain an even better understanding of the outbreaks at the time. This is the aim of the present study, which takes the canton and the city of Zurich as a case study to go into greater depth. We use newly digitized weekly aggregated data to re-model the 1918-1920 pandemic in Zurich and investigate how different parameters of mortality and disease burden correspond, how transmissibility changed during the different waves of 1918-1920, and how public health interventions were associated with changes in these pandemic parameters.

## 2. Data and methods

### The canton and the city of Zurich around 1918

In 1918, the city of Zurich was the largest city in Switzerland with a population of 211,850. The canton of Zurich was the second largest in terms of population (531,800 inhabitants). A look at the historical context of the 1918 pandemic shows that life in Switzerland at the time of the outbreak was dominated by the burdens of the war years: inflation, food rationing and, especially in cities like Zurich, and housing shortages. Real wages fell by around 25-30% between 1914 and 1918. The cost of living rose by 130%, in urban centres by as much as 150%.^19^ After rising steadily in the first and second phases of the war, inflation worsened in the third phase from 1917. This caused the poverty line to rise dramatically, and the situation peaked in November 1918.^29^

The tense social situation was further exacerbated by political issues that had arisen since the beginning of the war. The tensions culminated in the national general strike in November 1918 ^30^: Heralded by several smaller strikes, the general strike was called on 7 November 1918. Politically, the gathering of troops in central locations throughout Switzerland in the context of the influenza epidemic (many soldiers were suffering from the flu and spreading the disease to the civilian population) led to heated discussions ^30,31^. At the peak of the strike, which began on 9 November, some 8,000 soldiers faced tens of thousands of strikers in Zurich. Despite the tense atmosphere, the strike remained largely peaceful and was called off on 14 November 1918.

### The 1918 influenza pandemic in Switzerland and Zurich

There is currently no recent work on the 1918 influenza pandemic for the canton or city of Zurich; the last narrative study dates from 1968 ^32^. Cursory cantonal comparisons show that Zurich and the other cantons of eastern Switzerland were less affected than the Western cantons by the 1918 summer wave in terms of mortality ^20,33^. The reasons for this are still unclear. After a stabilization of the situation in September 1918, a very strong and long autumn and winter wave followed from October 1918 to the beginning of 1919, which affected the whole country equally.

If historical statistical surveys from shortly after the pandemic are followed, a total of 95,601 cases of flu were reported by physicians for the canton of Zurich in the year 1918 ^33^. This corresponds to 17.1 cases per 100 inhabitants, which is slightly higher than the Swiss average of 16.6 cases per 100 inhabitants. In 1918, 2370 deaths from flu were reported in the canton of Zurich (4.5 deaths per 1000 inhabitants, below the Swiss average of 5.5), 60.0% of whom were men. In the city of Zurich, 920 deaths (57.0% men) from flu were reported in 1918, or 4.3 deaths per 1,000 inhabitants. According to the Statistical Yearbook of the City of Zurich, the sexes and age groups were affected differently by flu mortality in 1918 as young adults and especially men were disproportionately affected (Supplementary Figure S2). A breakdown of flu deaths by sex and age by month in 1918 shows that the summer wave 1918 mainly affected young men aged 20-29, while the second wave also affected both sexes and other age groups (Supplementary Figure S3).

The start of the outbreak in Zurich can be approximately dated to mid-June 1918 ^32^: It is known that between 17 and 19 June 1918 about 30% of the workers in a factory in Zurich-Aussersihl came down with the flu. On 24 June 1918, similar illnesses were reported in a factory in Zurich-Albisrieden, and at the same time there were reports of individual cases of illness in Winterthur. In addition, most of the workers at a large industrial company in Zurich-Oerlikon fell ill, and the reports from all sides became numerous.

### Public health interventions in Zurich during the pandemic

A detailed schedule of official measures at various administrative levels can be found in Supplementary Figure S5. Responsibility for public health interventions in Switzerland in 1918 was with the 25 cantons (= member states) and not with the federal government. Even in 1918, the latter hardly intervened centrally, with the exception of a federal decree of 18 July 1918 authorising the cantons and municipalities to prohibit events that could lead to mass gatherings, and the nationwide obligation to report new influenza cases, which was introduced on 11 October 1918 (and thus much later than many cantons, including Zurich, which had already done so in July 1918).

The first intervention by the cantonal authorities took place on 25 July 1918, a few weeks after the epidemic had reached the canton and city of Zurich in late June or early July 1918. The package of measures that came into force on 25 July 1918 included an obligation for doctors to report cases of influenza and a ban on theatrical performances, cinematographs, concerts, public meetings, public festivals, preaching in churches and club halls. Only five days later, on 30 July 1918, the city of Zurich tightened these cantonal measures. In addition, meetings and events of any kind were banned in the city, pharmacies’ opening hours were extended, the number of seats in public houses and the number of passengers in trams were reduced, and the summer holidays of the public schools (which had begun at the beginning of July) were extended until 24 August 1918.

In August 1918, after the first wave had subsided, the canton of Zurich relaxed its measures in two stages: From 23 August 1918, meetings were permitted again in selected districts (Zurich, Affoltern, Horgen, Meilen and Dielsdorf) and public schools were reopened after the prolonged summer holidays. From 28 August 1918, certain celebrations were permitted again, but dances remained banned. After a relatively quiet period in September, the number of cases increased again at the beginning of October 1918. Emergency flu hospital units were set up, including in the prestigious Tonhalle concert hall (Supplementary Figure S1). The authorities reacted as follows: On 11 October 1918, the same restrictions were imposed as on 25 July 1918. In addition, public schools were closed for two months until 11 December 1918. The general strike of November 1918 caused a further increase in the number of flu cases at the peak of the second wave. The ban on assembly was ignored and tens of thousands of strikers and army troops gathered in the canton’s central squares. From the beginning of December 1918, the situation began to ease, the ban on meetings was gradually relaxed, schools were reopened, etc. From 14 December 1918, public meetings, celebrations of all kinds and church festivals were again permitted, although dancing and singing were still forbidden. However, the Zurich Health Department continued to advise against public Christmas celebrations. On 23 May 1919, all federal decrees to combat the influenza epidemic were repealed; only the obligation to report cases of influenza remained.

### Data sources

For this study, we digitised and analysed for the first time the following historical demographic and epidemiological data for the city and canton of Zurich:

#### a) Weekly new cases, deaths, and hospitalizations, 1910-1920

Since the end of the 19th century, the Federal Health Office has published a weekly bulletin on vital statistics, newly reported cases of notifiable infectious diseases and hospitalisations. We have digitised and transcribed the following weekly series for the period January 1910 to December 1920.

- For the city of Zurich: Deaths from all causes. These figures refer to both the resident and non-resident population. The quality of these historical vital statistics is considered in the literature to be very good, with incompleteness and migration no longer a problem compared to earlier years ^34^. However, death figures by age, sex and cause were not available on a weekly basis.

- For the city and canton of Zurich: New cases of influenza reported by physicians. This series begins with the introduction of mandatory reporting of influenza in the canton of Zurich in mid-July 1918. The authorities at the time estimated that these figures were probably underestimated 3-4-fold due to unreported mild cases ^33^.

- For the canton of Zurich: hospitalisations due to the category “Infectious diseases in general and influenza in particular”.

This weekly bulletin data was selectively supplemented by other weekly data by the authorities that we found in the course of our archive research, e.g. on the difference between the time of notification and the time of illness or influenza as a cause of death.

#### b) Monthly sick leave in a large Zurich-based company, 1918-1920

Swiss Re is a reinsurance company founded in Zurich in 1863. In January 1919, Swiss Re counted 406 employees (248 men and 158 women). From the Swiss Re Historical Archives SRHA (ID Number 10.120 259) we transcribed aggregated monthly sick days and total working and absences days by sex for the period between January 1918 and December 1920.

### Statistical methods

The weekly incidence, hospitalization and all-cause mortality rates of the canton and city of Zurich were calculated per 10,000 inhabitants (taken from census data). The weekly population was estimated by linear interpolation between the annual population figures.

The weekly all-cause excess mortality was estimated by using a Bayesian approach (INLA). The number of deaths was modelled using a Poisson distribution, with the weekly population considered as an offset. The latent time trend was integrated as a random walk of order 1, and seasonality was modelled using a seasonal variation model with a periodicity of s=12, as described in INLA ^35–37^. The calculations were based on data from the last five years of the respective year and weeks, excluding pandemic years with high mortality (1918 and 1920). After fitting the model, we drew 1000 samples from the posterior distribution (i.e. the expected number of deaths). Excess rates were then computed by deducting the expected values from the observed ones. Subsequently, the excess mortality rates were presented as percentages.

The relative incidence rate (RIRR) was calculated according to Staub et al ^38^. First, the incidence rate ratio (IRR) was computed for each calendar week i, obtained by dividing the incidence rate of week i by the incidence rate of the preceding week i – 1. Following that, we calculated the RIRR by dividing the IRR of week i, by the IRR of the preceding week. An RIRR greater than 1 indicates an acceleration in the growth of the epidemic, while an RIRR less than 1 indicates a slowdown (equivalent to the second derivative).

The effective reproduction number *R_e_* was estimated using a negative binomial generalized linear model accounting for overdispersion. The last 3 weeks (21 days) before the respective week are used for estimating the epidemic growth rate which can be used to compute *R_e_*. To this end, we assumed a gamma-distributed generation time with a mean of 3 days and a standard deviation of 1 day ^17,39,40^.

All statistical analyses were performed using R Version 4.2.2 ^41^. R-INLA ^35^ was used to estimate the excess mortality and “tidyverse” ^42^ was used to process the data and create the remaining figures.

## 3. Results

The course of the pandemic based on the various parameters is shown in **Figure 1**. From the end of June or beginning of July 1918, the various parameters begin to increase for the first wave in the summer of 1918. Because the mandatory reporting of influenza in the canton of Zurich was not introduced until July 25, 1918, the beginning of the first wave is less well represented by the incidence figures. The various parameters for the first wave agree well in that incidence, hospitalizations and sickness days go through the wave in July to August 1918, but deaths and excess mortality, on the other hand, only show a moderate increase at the same time. The effective reproduction number *R_e_* based on incidence and hospitalizations is around 1.7 (**Table 1**). The first package of public health measures in calendar week 29 was not associated with any significant change in the course of the epidemic, but the tightening of the measures in week 30 (July 30, 1918) shows an association with the decrease in epidemic growth around 1-2 weeks later (RIRR 0.25 (95%CI 0.22-0.29)) (**Table 2**).

**Figure 1:**
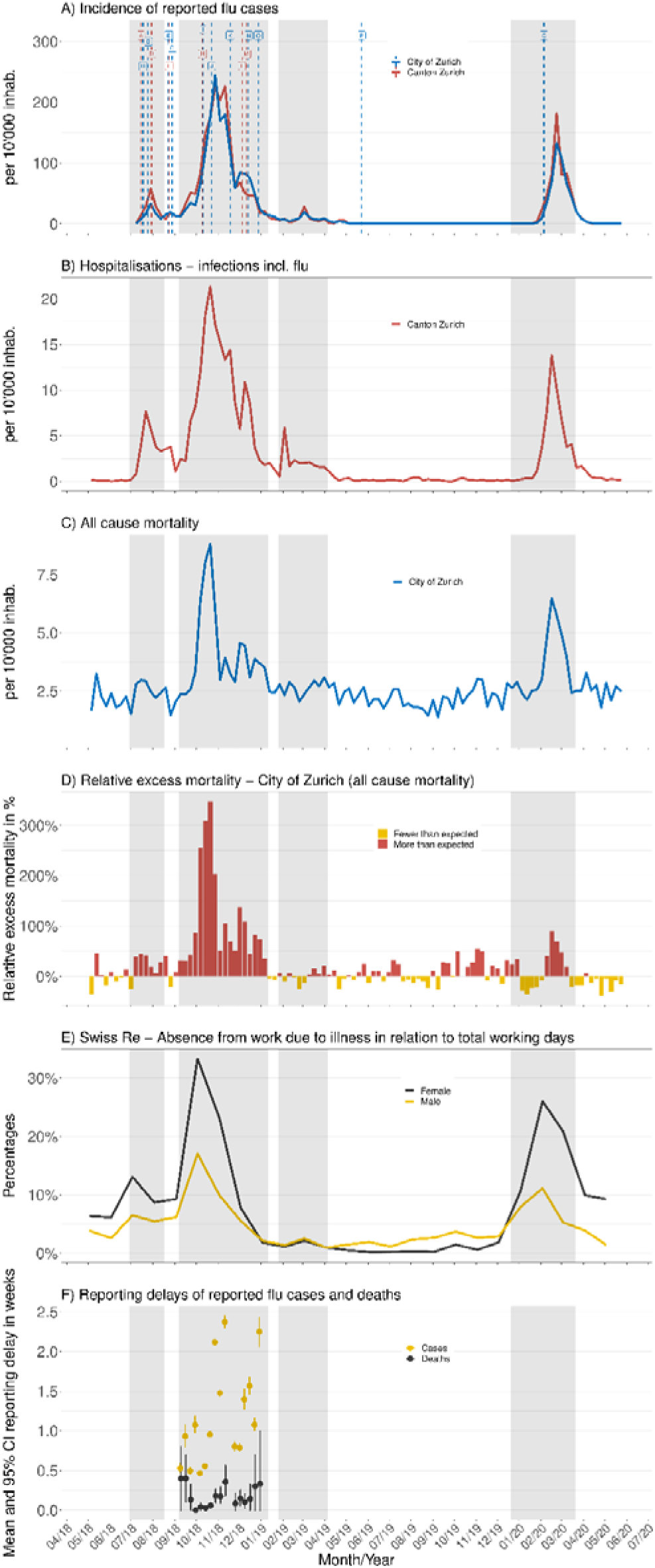
A-D) Weekly numbers of reported cases, hospitalisations, all-cause deaths, and excess mortality during the influenza pandemic in Switzerland in 1918-1920; E) Monthly sickness absences Swiss Re; F) Reporting delays in Fall 1918. The letters in A) lead to the detailed interventions as outlined in Supplementary Figure S5).

**Figure 2:**
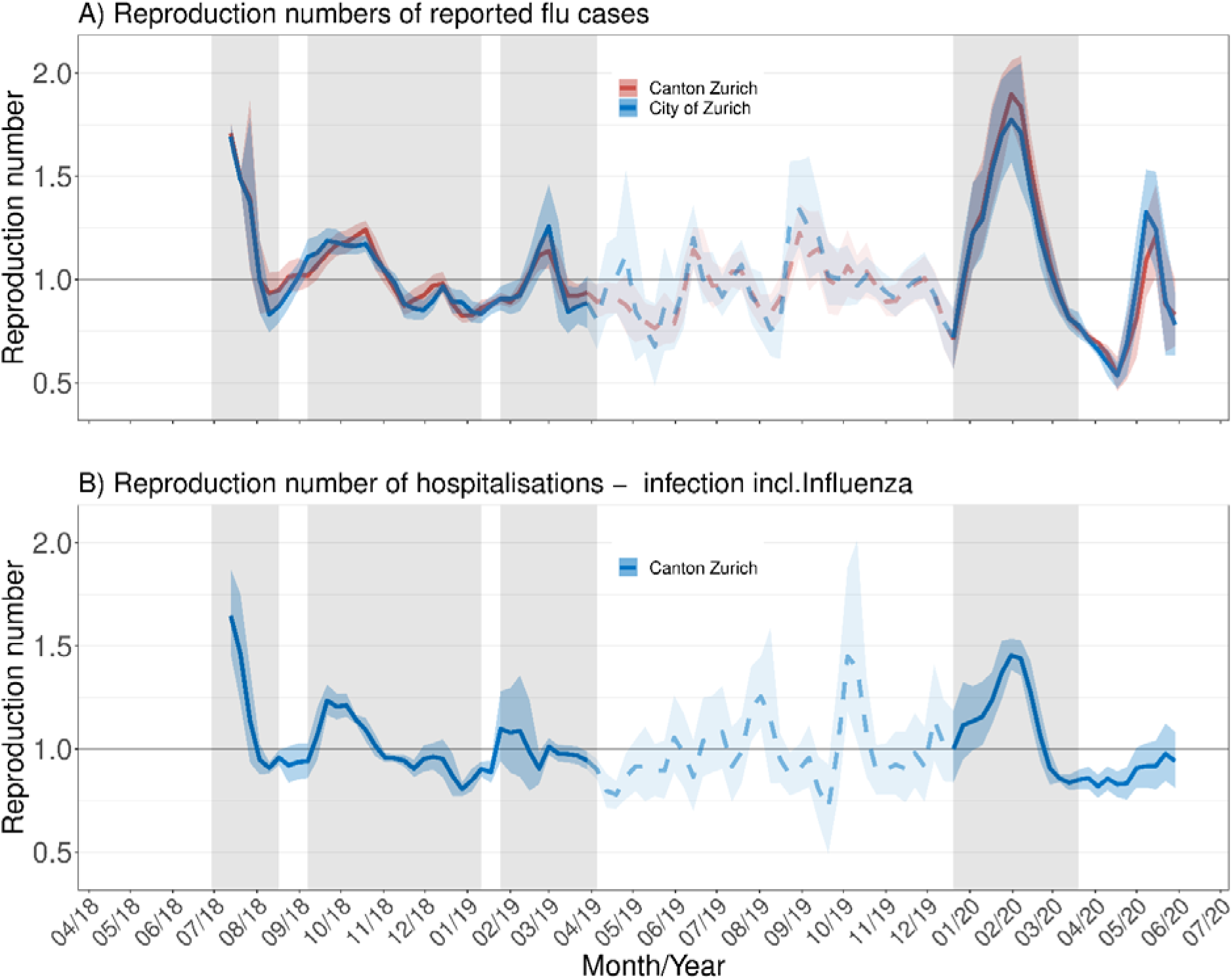
Temporal changes of the effective reproduction number *R_e_* for A) reported cases and B) hospitalizations due to infections including influenza.

**Table 1:** Effective reproduction number *R_e_* for the 4 waves 1918-1920 and different pandemic parameters.

| Wave | Year | Week | Cases Canton | Cases City | Hosp. Canton |
| --- | --- | --- | --- | --- | --- |
| 1 | 1918 | 28 | 1.71 [1.65-1.77] | 1.69 [1.63-1.76] | 1.65 [1.46-1.87] |
| 2 (start) | 1918 | 38 | 1.12 [1.07-1.18] | 1.19 [1.13-1.25] | 1.24 [1.16-1.31] |
| 2 (strike) | 1918 | 40 | 1.18 [1.13-1.24] | 1.16 [1.12-1.21] | 1.21 [1.16-1.27] |
| 3 | 1919 | 9 | 1.14 [1.05-1.22] | 1.26 [1.08-1.46] | 1.01 [0.97-1.05] |
| 4 | 1920 | 5 | 1.90 [1.75-2.06] | 1.77 [1.57-2.02] | 1.45 [1.38-1.54] |

**Table 2:** Association between selected non-pharmaceutical interventions and epidemic growth, as indicated by Relative incidence rate ratios (RIRR). An RIRR >1 indicates acceleration of epidemic growth 1-2 weeks after an intervention or event, whereas an RIRR <1 indicates deceleration (details on the interventions can be found in Supplementary Figure S5).

| ID (Fig. 2A) | Intervention/Event | Enactment Wk | Impact Wk | Cases | Cases previous Wk | Cases 2 Wk before | RIRR (95% CI) |
| --- | --- | --- | --- | --- | --- | --- | --- |
| A + B | Reporting obligation, etc. | 29 | 30 | 1784 | 892 | 446 | 1.00 (0.85-1.18) |
| C | School closures & assembly bans | 30 | 31 | 892 | 1784 | 892 | 0.25 (0.22-0.29) |
| E+F+ G | Relaxations & school openings | 34-35 | 36 | 647 | 711 | 1036 | 1.33 (1.11-1.58) |
| H + I | School closures & assembly bans | 41 | 42 | 9436 | 6319 | 3495 | 0.83 (0.78-0.88) |
| K | General strike November 1918 | 45-46 | 48 | 4523 | 3121 | 6358 | 2.95 (2.73-3.20) |
| L | First relaxations | 49 | 51 | 2642 | 3988 | 4431 | 0.74 (0.68-0.80) |
| M + N | School openings & further relaxations | 50 | 52 | 1329 | 2642 | 3988 | 0.76 (0.69-0.84) |

After epidemic activity fell to a low level at the end of August and especially in September 1918, the bans on gatherings were relaxed and schools reopened. The lifting of these measures in calendar weeks 34 and 35 was associated with a renewed increase in the number of cases from the beginning of October 1918 (RIRR 1.33 (95%CI 1.11-1.58)) and the beginning of the second and strongest wave (**Table 2**). The effective reproduction numbers are slightly lower for the second wave (around 1.2, see **Table 1**). This time, in addition to the incidence and hospitalizations, all-cause deaths and excess mortality are also increasing sharply. In certain weeks at the end of October 1918, the excess mortality rate for all causes reached around 300% compared to the expected weekly values based on previous years. At the same time in October 1918, Swiss-Re’s female employees missed around 30% of all regular working days due to illness. After this unprecedented increase in pandemic parameters in October 1918, the authorities had to react again and in calendar week 41 issued the same package of measures as in the summer of 1918 (bans on gatherings and school closures). This was in turn associated with a decrease in epidemic growth (RIRR 0.83 (95%CI 0.78-0.88)) 1-2 weeks later.

After the second wave was successfully contained for the first time at the end of October, the socio-politically motivated national strike took place in all major places in Switzerland, including Zurich, in early to mid-November (calendar weeks 45 and 46). It lasted several days, mostly peacefully, and the thousands of strikers were confronted by at least as many soldiers, many of whom had the flu and were crowded together in schools and other facilities. One to two weeks later, the period of the national strike was also associated with a resurgence of cases (RIRR 2.95 (2.73-3.20)) (**Table 2**), and the otherwise already powerful second wave was temporarily extended. All pandemic parameters show this resurgence of the pandemic wave in the second half of November (the reproductive numbers are again around 1.2 (**Table 1**)).

For the second wave, thanks to detailed historical sources for deaths and newly reported cases, the date of illness or death can be compared with the date of notification and registration (**Figure 1F**). In the case of deaths, even at the peak of the second wave, the average of reporting delay was rather <0.5 weeks, so there was no major delay in reporting deaths. In contrast, the reporting and registration delay for new cases was usually 0.5-1.0 weeks and thus a bit longer than for deaths. The influence of the national strike is also very clear in the case of the reporting delays of cases when new flu illnesses had an average reporting delay of 1.5-2.5 weeks in November 1918.

The second wave gradually levelled off in December, and the measures were successively lifted until the end of 1918, without this leading to a resurgence in the number of cases. In March 1919, certain pandemic parameters (cases, hospitalisations, sick days) show a very slight increase, but a clear wave is not visible in Spring 1919. On the other hand, a later wave of the pandemic occurred one year later in January to March 1920, which again had a marked impact on all pandemic parameters (*R_e_* between 1.5 and 1.9 (**Table 1**)), including excess mortality. All the pandemic parameters analysed show higher levels in this later wave (in which there was hardly any government intervention) than in the first pandemic wave in the summer of 1918 (**Figure 1**).

## 4. Discussion

We have re-modelled the course of the 1918-1920 influenza pandemic in Zurich using various epidemic/pandemic time series (incidence, hospitalisations, mortality and sickness absences from work). In general, the various parameters show a good temporal correspondence, but differences in their expression can also be observed. The first wave in the summer of 1918 did lead to illness, absence from work and hospitalisations, but to a lesser extent to increased mortality. In contrast, the second, longest and strongest wave in the autumn/winter of 1918 also led to greatly increased (excess) mortality in addition to the burden of illness. While there was hardly a noticeable wave in Zurich in spring 1919, the later wave in the first months of 1920 was again associated with an increase in all pandemic parameters. This later 1920s wave was even stronger overall in Zurich than the first wave in the summer of 1918. Furthermore, we can see that public health measures such as bans on gatherings and school closures were associated with a decrease in the course of the pandemic, while the lifting or non-compliance with these measures was associated with an increase of reported cases.

With regard to the estimated transmission dynamics of the various pandemic waves 1918-1920, there are a number of other studies in the literature, including some from Switzerland, with which our results can be compared. For example, a study on comparable incidence figures for the canton of Bern found very similar reproduction numbers for the second wave in autumn/winter 1919 (1.2 vs 1.3) ^38^. The initial wave in the summer of 1918 had a higher reproduction number in both Bern and Zurich than the second autumn/winter wave. However, the reproduction number in Bern was even higher for the first wave than in Zürich (2.3 vs. 1.7), which could be due to the fact that Bern had introduced compulsory notification of new influenza cases around 10 days before Zurich, and therefore the beginning of the summer wave is better reflected in the case numbers. On the other hand, the summer wave was also stronger west of the Central Plateau, which could be another reason. In Bern, the small wave in the spring of 1919 was much more pronounced than in Zurich, but the later wave at the beginning of 1920 was equally strong in both Bern and Zurich ^27^. Reproduction numbers were already estimated almost 20 years ago for the canton of Geneva, where a higher reproduction number (3.8) was estimated for the second wave than for the first wave (1.5) in 1918 ^25,26,43^. It remains to be clarified whether the underlying different data basis for Geneva (daily hospitalisations), different methods (a compartmental model with inclusion of underreporting), different assumptions of the generation time or possibly actual differences in the course of the epidemic are responsible for these differences. In an international comparison, the reproduction number of the second wave in Zurich (1.2) is roughly comparable with data from Copenhagen, Gothenburg or Oslo (1.2-1.5) ^44,45^. However, the reproduction rate of the first wave in Zurich (1.7) is rather low not only in comparison with Bern (2.3), but also in international comparison with the Scandinavian cities (2.0-4.8, see above for possible reasons) ^44^.

Before Covid-19, the influenza pandemic of 1918-1920 was the last pandemic that was distinctly multiwave, lasting 2-3 years. In addition to many important differences worth emphasising between COVID-19 and the 1918-1920 influenza pandemic (different virus, different living context, etc.), one of the similarities was probably that several crises overlapped also in the period 1918-1920. The pandemic coincided with the end of the First World War, which had also left its mark on people’s nutritional status in Switzerland, which was not directly involved in the war, particularly from 1917 onwards. The fact that people were rather weakened in 1917 and 1918 could have contributed to the severity of the pandemic ^46^. In addition, there had already been socio-political labour conflicts for several years, which culminated in the general strike in November 1918 ^47^. We confirm also for Zurich (the same has already been shown for Bern ^38^) that the mass gatherings associated with the strike (especially of troops in buildings in the city) were associated with a resurgence of the epidemic during the second wave in the autumn/winter of 1918.

A remarkable aspect of our Zurich case study is the strong later wave in the first months of 1920. All pandemic parameters analysed, mortality and disease burden, show a stronger wave for Zurich in 1920 than the first pandemic wave in the summer of 1918. On the one hand, in comparison with other Swiss cantons and cities, we confirm for Zurich the so-called Eastern Switzerland pandemic pattern already described earlier on the basis of mortality figures ^19^, with a less pronounced first summer wave in 1918 compared to Western Switzerland, for example, but we broaden the Swiss horizon to include the obviously important 1920 wave, which has so far received too little attention ^27^. On the other hand, we also confirm a handful of studies on other countries that have already pointed to the relevance of this later wave ^48–50^. The exact reasons and possible explanations for this strong later wave still need to be clarified: There are initial signs that the virus has changed genetically during the pandemic ^51^, and that reinfections and cross-protection between waves therefore play a role ^52,53^. It also remains to be clarified to what extent the 1920 wave is still part of the pandemic waves or whether it should be considered a seasonal wave, which usually occurred every two winters in Switzerland from the 1920s onwards ^27^. Future investigations into the pattern of age groups affected by mortality in the 1920 wave and genetic changes in the virus may provide answers in the coming years.

Previous knowledge of the different patterns of the 1918-1920 influenza pandemic in Switzerland is based on descriptive death figures by month and canton, which were compiled and described in the 1990s. Due to the barely increased mortality figures in the summer of 1918, it was assumed at the time that, in contrast to western Switzerland, there was hardly any notable summer wave in eastern Switzerland in July and August 1918. On the one hand, we confirm this eastern Swiss mortality pattern for Zurich in the summer of 1918 with our weekly mortality figures. On the other hand, we also show that there was indeed a summer wave in Zurich, as indicated by the disease parameters, but it did not result in increased mortality as in western Switzerland. The exact reasons for these regionally differing patterns still need to be clarified. One factor could be that the summer holidays in schools were timed differently depending on the regions of Switzerland: while schools in eastern Switzerland were not yet open again at the start of the summer wave and the authorities did not have to close the schools but simply extend the holidays, the holidays in western Switzerland ended earlier and children were already back in school at the start of the pandemic before they had to be closed again by the authorities. This could have been an important factor in the spread of the pandemic wave.

The strength and duration of non-pharmaceutical interventions such as school closures and assembly restrictions in the context of the 1918-1920 influenza pandemic have been shown several times to have had an impact on mortality ^54–56^. The results of our Zurich case study are in line with an earlier study on the incidence of flu in 1918 in the canton of Bern and supports the notion that school closures and assembly bans were associated with slowing down epidemic growth and breaking waves ^38^. The fact that shortly after the peak of the second wave, mass gatherings in the context of the national strike caused the pandemic wave to rise again in both Zurich and Bern also indicates how important it is for the population to comply with non-pharmaceutical measures. This could also have been an important factor at the beginning of October 1918, i.e. between the waves and at the beginning of the second wave. A long article in the Neue Zürcher Nachrichten (one of the largest daily newspapers) on 8 October 1918 written by the official city physician at the time, should be mentioned here as an anecdote. In this article he writes to the public: “The flare-up of the epidemic is primarily favoured by the carelessness of the public, bordering on recklessness, who, having barely escaped the danger, throw all well-meant advice to the wind, whether out of comfort or out of selfishness and pleasure-seeking.” And further down: “A new wave of the flu epidemic is coming … hundreds of people may have to pay for their carelessness with their lives tomorrow.” Finally, he calls out: “It must therefore be called the duty of every individual to contribute to the containment of the epidemic in his own place, putting aside his own self-interest, and by conscientiously following all that is required.”

To our knowledge, there are not many studies on sickness-related absences from work during the 1918-1920 pandemic ^57^. Our data shows that medium-sized and larger companies were also considerably affected. It must be left to follow-up studies to analyse these effects on companies more comprehensively. What is clear, however, is that sickness-related absences from work were already an area of concern during the 1890s pandemic, as has already been shown for postal and railway companies in Switzerland ^58,59^.

Our study has various limitations: First, with the selected parameters we only depict the immediate disease and mortality consequences of the 1918-20 pandemic. There are also medium- and long-term consequences of the flu illnesses, for example in terms of post-viral symptoms and later illnesses ^60–63^. Such patterns have also been documented for *in utero* exposure to the 1918-1920 influenza pandemic ^64^. For an even more holistic view, such aspects should also be included. Secondly, the incidence numbers in particular are either underestimated or overestimated, despite or precisely because of the reporting obligation. Added to this is the reporting delay at the peak of the second wave. This must be taken into account in the interpretation. However, our analysis shows that the incidence numbers provide a solid picture of the course of the pandemic in comparison with the other parameters, particularly in terms of timing. Thirdly, we analysed aggregated data in our paper, which meant that we did not have access to individual information on age, sex or occupation that would provide additional insights. Fourthly, this study cannot address whether the results of this Zurich case study can be generalised to a different context. Given that the pandemic experience could vary regionally in Switzerland ^19,27^, we assume that the Zurich results can at least be applied to Eastern Switzerland, but this still needs to be investigated. Caution must also be used regarding the generalisability of the results from the 1918-1920 pandemic to other and possibly future pandemics, as certain experiences are certainly similar, but other pandemics always show differences and take place in a different living context ^65^. Finally, our study shows associations, and causality cannot necessarily be inferred from this.

## 5. Conclusion

With our study, we point out how important it is not to focus exclusively on one pandemic parameter (in the literature so far mostly mortality), but to analyse the disease burden of a pandemic more comprehensively. Incidence, hospitalisations and absences from work all represent related but slightly different aspects of the overall disease burden, and nuances between the parameters represent the dynamics during a pandemic. This is especially true when a pandemic is multi-wave. And the last pre-COVID-19 pandemic that was distinctly multi-wave was the 1918-1920 influenza pandemic. Certain challenges explicitly associated with multi-wave may well be similar and comparable between pandemics. Experiences from the 1918-1920 pandemic in this regard have been forgotten over time. We want to counteract this forgetting of such potentially valuable past experiences with our contribution, also with regard to similar challenges in the future.

## Author statements

## Data Availability

All data produced and used are available online at https://zenodo.org/records/7986584

## Acknowledgements

The authors would like to thank Inga Birkhölzer and Julia Simola for their help in transcribing the historical data, Swiss Re Historical Archives (SRHA) for access to their archives, and Wibke Weber, Maryam Kordi, Gian Hedinger, Peter Jüni, Marcel Zwahlen, Olivia Keiser, and Joël Floris for previous/ongoing collaborations and helpful comments.

## Author contributions

Conceptualization: EZ, KS, CA, KM

Data curation: EZ, KM, PM

Methodology: KM

Data analysis: KM

Funding acquisition: KS, FR

Supervision: KS, VS

Writing – first draft: EZ, KS

Writing – review & editing: All authors.

## Ethics committee approval

Ethics approval was not required for the reuse of these publicly available and aggregated data.

## Reproducible Research Statement

Protocol: Not available.

Statistical code: https://github.com/KaMatthes/Influenza_Zurich_1918_1920 Data: https://zenodo.org/records/7986584

## Supplementary Material

**Supplementary Figure S1:**
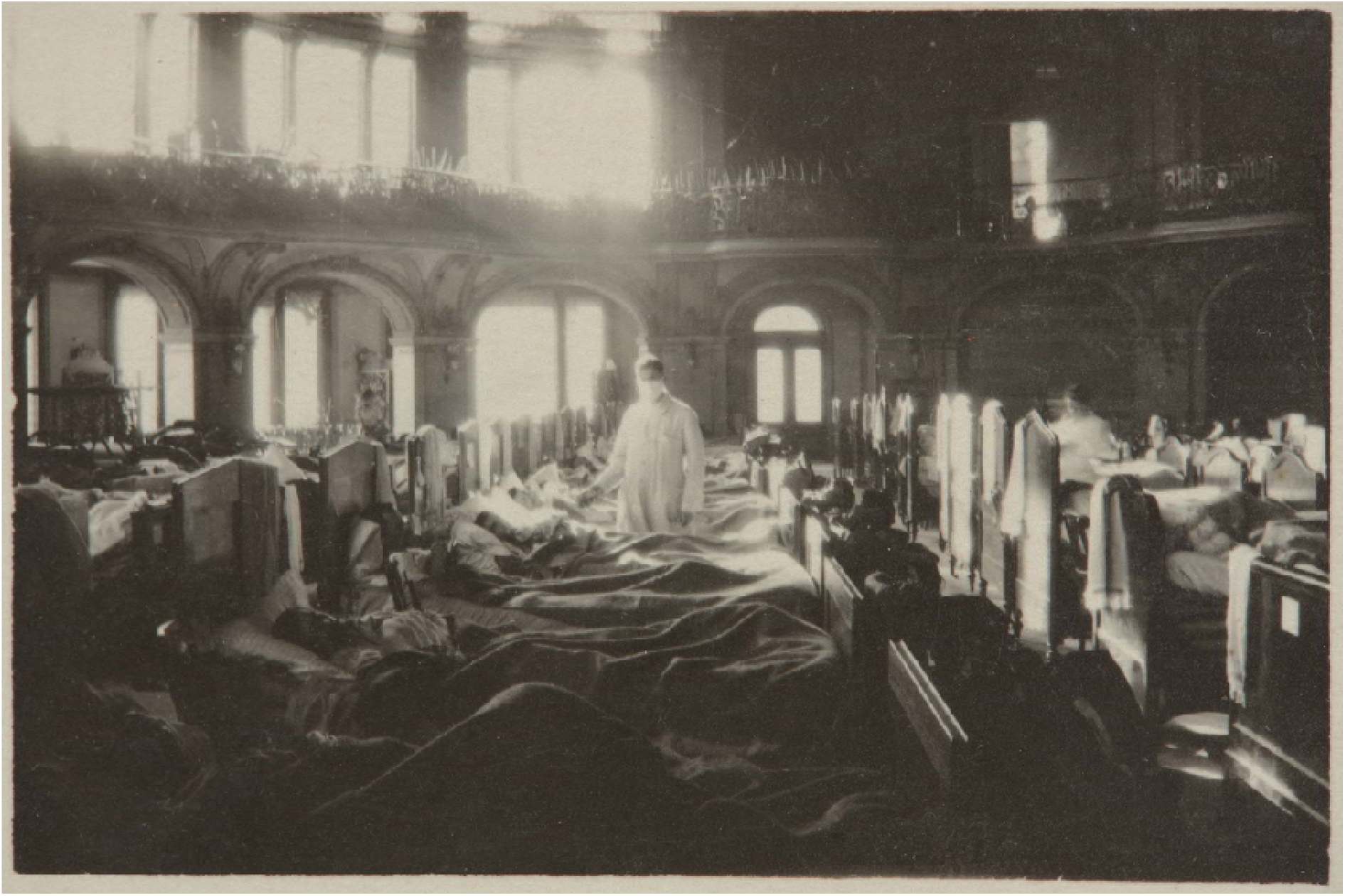
Contemporary photograph of the flu emergency hospital in the Tonhalle in the city of Zurich, in November 1918 (Source: Schweizerisches Nationalmuseum, Inventar-Nummer LM-102737.46).

**Supplementary Figure S2:**
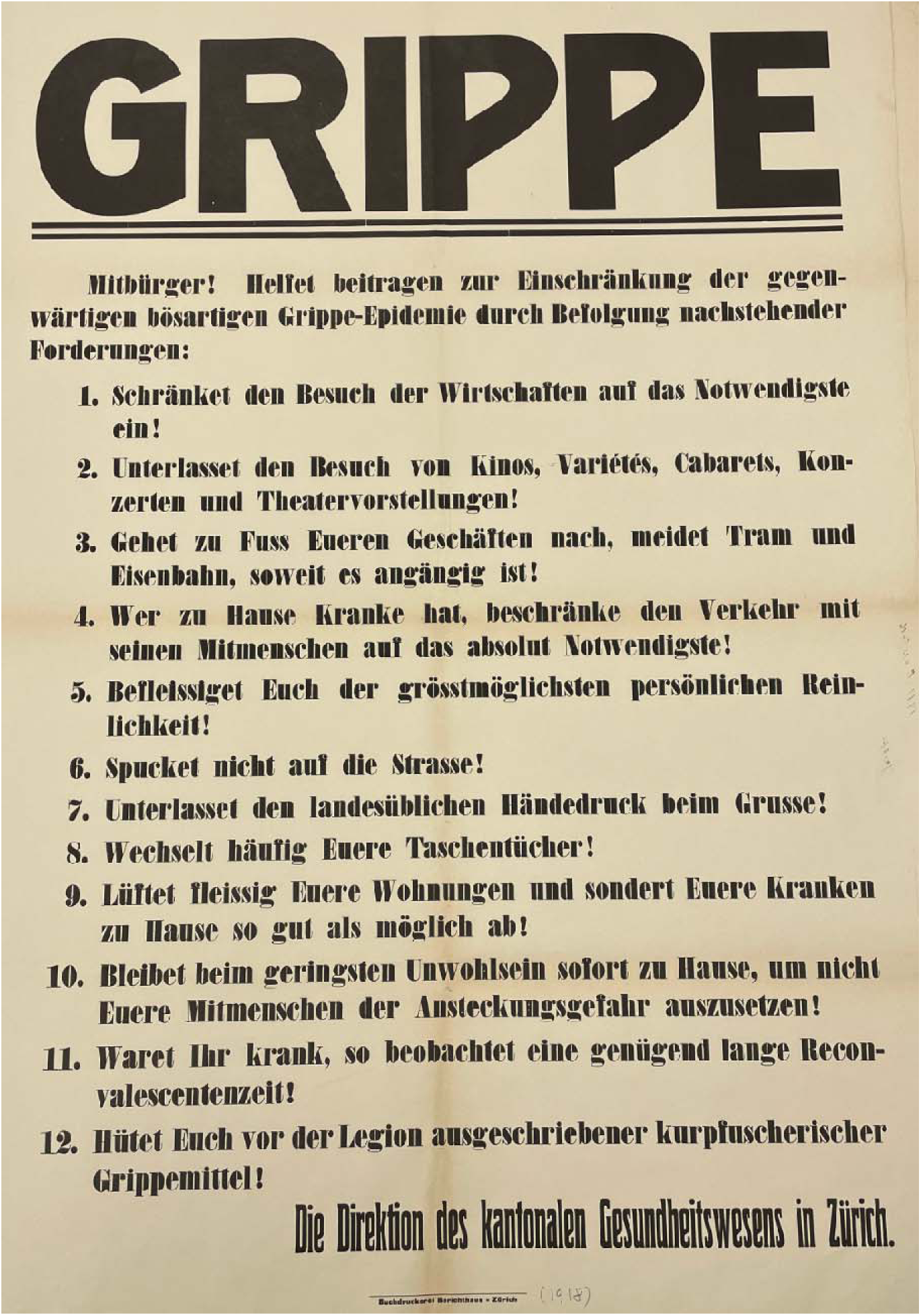
Poster of the Zurich health authorities with behavioral instructions from the fall of 1918 (source: State Archives of the Canton of Zurich).

**Supplementary Figure S3:**
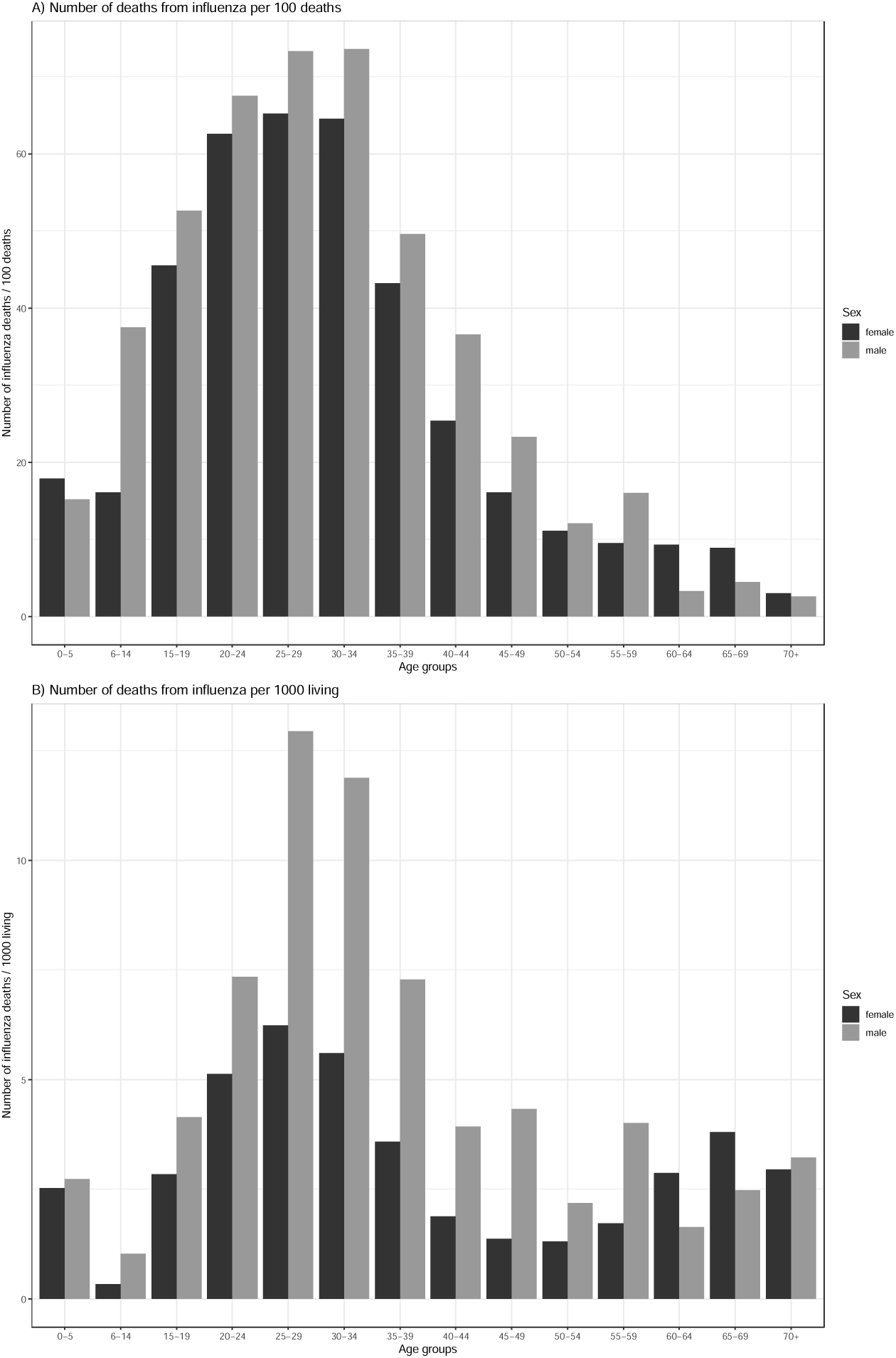
From the statistical yearbook of the City of Zurich: Visualization of aggregated numbers of flu deaths by age group and sex for the year 1918, A) in relation to all deaths and B) in relation to the population.

**Supplementary Figure S4:**
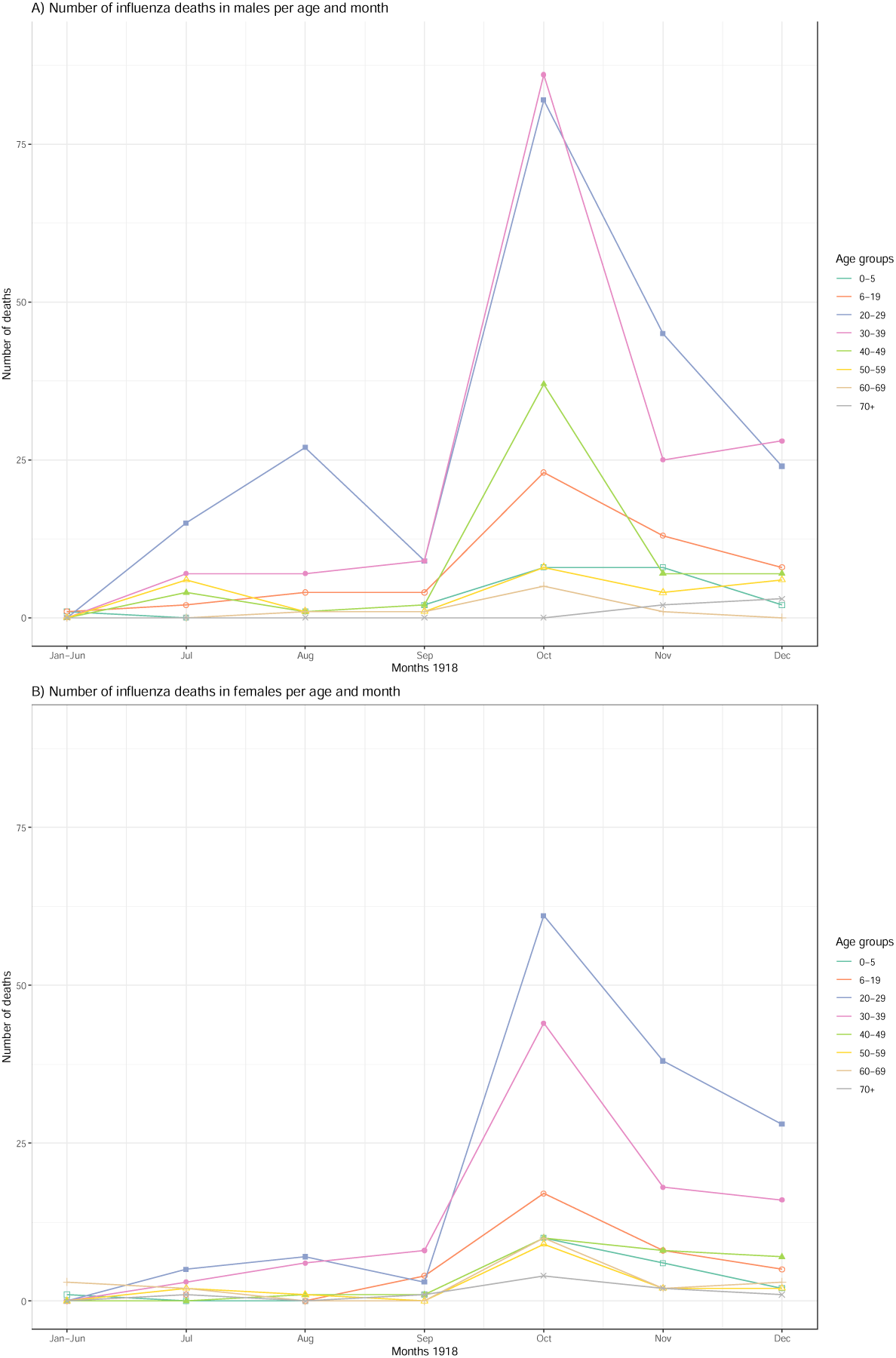
From the statistical yearbook of the City of Zurich: Visualization of aggregated numbers of flu deaths by age group, sex and month for the year 1918, A) males and B) females.

**Supplementary Figure S5:**
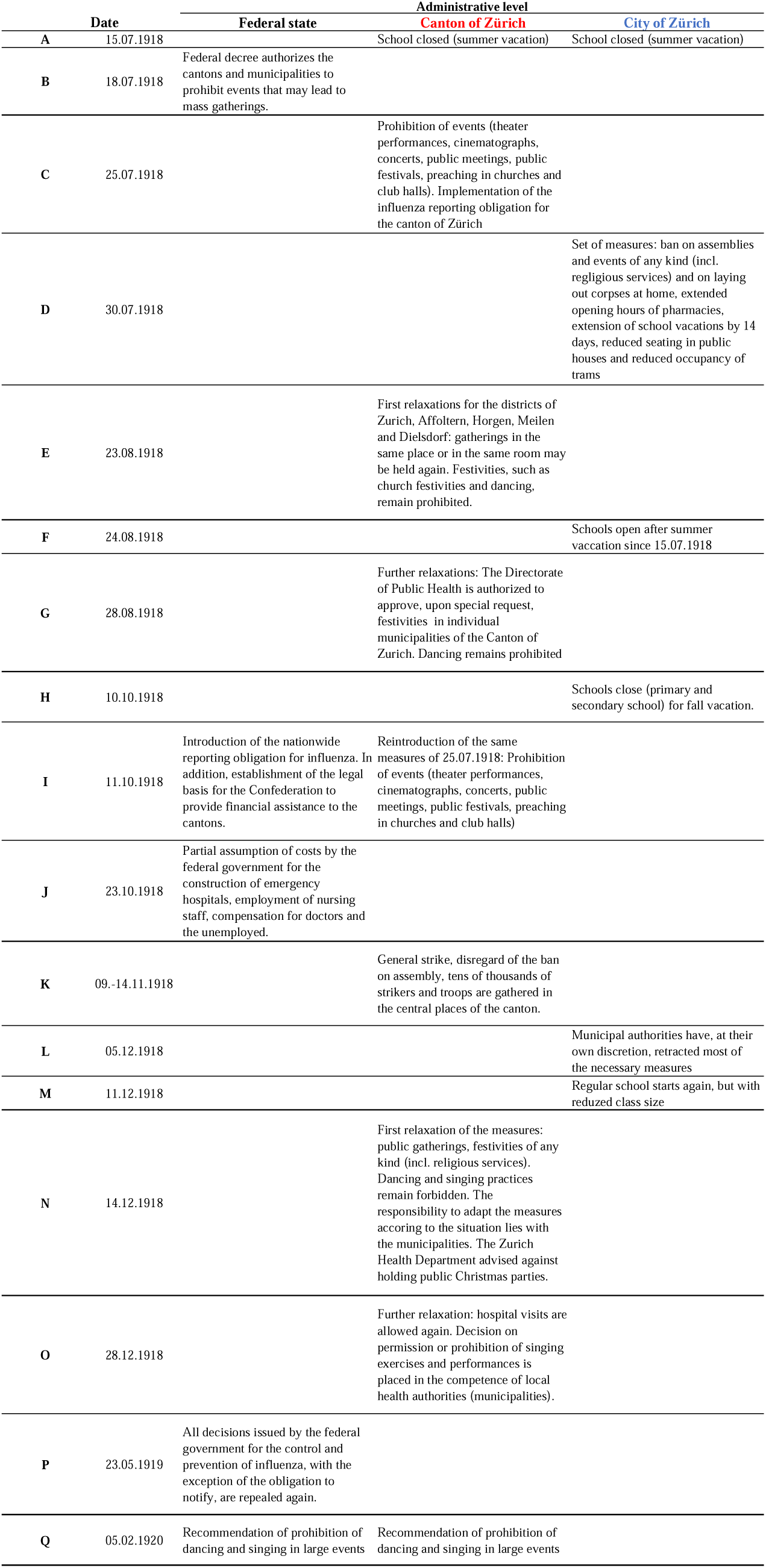
Detailed schedule of the official interventions at various administrative levels.

